# Development of an accessible gene expression bioinformatics pipeline to study driver mutations of colorectal cancer

**DOI:** 10.1101/2021.11.10.21266196

**Authors:** Lisa van den Driest, Caroline H Johnson, Nicholas JW Rattray, Zahra Rattray

## Abstract

Colorectal cancer (CRC) is a global cause of cancer-related mortality driven by genetic and environmental factors which influence therapeutic outcomes. The emergence of next-generation sequencing technologies enables the rapid and extensive collection and curation of genetic data for each cancer type into clinical gene expression biobanks.

In this study we used a combination of bioinformatics tools to investigate the expression patterns and prognostic significance of two genes, adenomatous polyposis coli (*APC*) and B-Raf proto-oncogene (*BRAF*), that are commonly dysregulated in colon cancer. Subsequently, we investigated the pathways and biomolecular effectors implicated in *APC* and *BRAF* function.

Our results show mutation types, frequency, anatomical location and differential expression patterns for *APC* and *BRAF* between colorectal tumour and matched healthy tissue. The prognostic values of *APC* and *BRAF* was investigated as a function of expression level in CRC and other cancer types.

In the era of precision medicine and with significant advancements in biobanking and data curation, there is significant scope to use existing clinical datasets for evaluating the role of mutational drivers in carcinogenesis. This offers the potential for studying combinations of less well-known genes and the discovery of novel biomarkers or studying the association between various effector proteins and pathways.

## Introduction

Colorectal cancer (CRC), also known as bowel cancer, is a type of cancer developed by uncontrolled cell growth in parts of the large intestine or the appendix and is a major cause of global morbidity and mortality. In 2020, CRC was ranked as the third most common cancer with 1,931,590 cases (10%) and the second highest mortality rate with 935,173 cases (9.4%) worldwide^1^. It is predicted that the number of new cases will rise to approximately 2.5 million in 2035^2^. Surgery and chemotherapy are routine standard of care interventions used to treat CRC. Novel improved treatment methods such as targeted therapies against genetic mutations and the rapid development of screening techniques such as (virtual) colonoscopy, sigmoidoscopy, stool-based tests and liquid biopsies have shown to detect the disease sooner and extend overall survival rates for patients with CRC. Overall, the 5-year survival rate for CRC is ∼64%.^3^

CRC is a heterogeneous disease driven by both genetic instability and environmental factors^5,6^. Major advancements in DNA sequencing technologies are resulting in a rapid expansion of biobanks and repositories for patient tumour biopsies. This significant wealth of data offers scientists the information to examine common gene mutations, their cellular pathways and their influence on tumour growth and development. A major challenge in the treatment of CRC is identifying the multiple variables that impact the emergence of metastases and their prognostic implications. Various pathways mediate the initiation, progression, and metastasis of CRC or those eligible in the activation of signalling cascades, contain ideal sites for targeted therapy. Therefore, identification and deeper understanding of common mutations in patients with CRC and their effects on cellular pathways are crucial to better understand and treat the disease^7^.

Studies related to the prevention, treatment and development of CRC use both animal models and cell cultures to mimic the cancer microenvironment for patients with CRC. A review study conducted in 2015 investigated the use of pre-clinical models in CRC by investigating current literature (477 relevant articles). Overall, each reported pre-clinical model showed limitations arising from a lack of spontaneous CRC development, or the requirement for a carcinogen for tumour induction in rodents. Moreover, there is significant inter-animal variability in the development of intestinal tumours for murine pre-clinical models.^8^ Alternatively, freely accessible clinical datasets contain a wealth of data to study expression patterns and evaluate the prognostic significance in of gene mutations in human CRC tissue, with the option to further investigate clinicopathologic parameters such as sex, anatomical location, stage and age.

Two genes important in CRC are the tumour suppressor gene adenomatous polyposis coli *(APC)* and the oncogene B-Raf proto-oncogene (*BRAF)*, implicated in CRC development^7,9^. The most common CRC activation pathway, which is responsible for 70-80% of all CRC diagnoses, is triggered due to inactivation of the tumour suppressor gene *APC*, which has an important role in the wnt signalling pathway, intercellular adhesion, cytoskeleton stabilisation, cell cycle regulation and apoptosis^10–12^. Therefore, truncating mutations of *APC* resulting in its inactivation are thought to promote tumorigenesis and adenoma formation, then subsequent mutations to KRAS and TP53 propagate the development of carcinomas, commonly found in left-sided colon cancers (LCCs). However, KRAS mutations are in higher frequency in right-sided colon cancers (RCC), together with mutations in the BRAF gene^13^. Recent epidemiologic studies have demonstrated that the anatomical location of CRC tumours impacts the overall patient survival, as patients with RCC were correlated to a poorer prognostic outcome^14–16^. Exploring the genetic and prognostic rationale of these gene mutations is important to identify new biomarkers for diagnosis, develop novel potential drug targets for treatment and explore new effective treatment combinations. In this study we intend to develop an integrated bioinformatics workflow that will blend orthogonal datasets and interrogate the two main cellular pathways as drivers of colon cancer aggressiveness in patient populations.

To-date, several studies have applied bioinformatics tools to investigate diagnostic and prognostic data, key pathways and important genes causing the progression of CRC. Some of these studies conduct a comparison between expression profiles of samples obtained from patients with CRC to those in normal healthy control^17–21^. Yang et al. used bioinformatic analysis to explore the overexpression or downregulation of microRNAs in CRC tissues and their role in tumorigenesis, invasion, and migration^22^. In another study online accessible databanks were used to study the chromobox family proteins and their role in CRC^23^. In this study we report the analysis of driver mutations (primarily *APC* and *BRAF*) in CRC using existing clinical datasets from a range of publicly available biobanks.

## Methods

### cBioPortal

(https://www.cbioportal.org/) is an open access resource for cancer genomics that was originally developed by the Memorial Sloan Kettering Cancer Center^24^. This source was used to investigate common gene mutations found in patients with CRC. A total of 7 studies (n=2,575) covering colorectal adenocarcinoma, metastatic colorectal cancer, colon adenocarcinoma, and colon cancer were explored, and a list of frequent mutated genes was generated^25–31^. Additionally, using the same seven studies, the *APC/BRAF* gene pairing was selected. Under the *‘cancer type summary’* tab, *‘cancer type’* was selected to generate a bar chart showing the alteration frequency in each cancer type.

### PrognoScan

(http://dna00.bio.kyutech.ac.jp/PrognoScan-cgi/PrognoScan.cgi) is a database for performing meta-analysis of the prognostic value of mutations occurring in cancer through incorporating gene expression studies from multiple sources such as the Gene Expression Omnibus (GEO) and reports from individual labs^32^. It relates gene expression data to prognostic outcomes, which enables the evaluation of potential biomarkers and their role in cancer prognosis. In this study PrognoScan was used to assess the correlation between *APC* and *BRAF* mRNA expression levels and patient overall survival (OS), disease free survival (DFS) and disease specific survival (DSS). Output generated from PrognoScan include *P*-values (Cox), hazard ratios and confidence intervals. *APC* and *BRAF* were independently entered in the PrognoScan website and the *P*-values (Cox), hazard ratios and confidence intervals were obtained for colorectal cancer studies. R Studio software in combination with the forestplot package were used to generate a forest plot from the values obtained via PrognoScan.

### Oncomine

(https://www.oncomine.org/resource/login.html) is a bioinformatic platform with an extensive collection of datasets^33^. Analysis of *APC/BRAF* mRNA expression pattern was performed selecting the following parameters: gene-*APC/BRAF*, differential analysis-cancer vs. normal analysis, cancer type-colorectal cancer, sample type-clinical specimen, data type-mRNA. Order by: Under-expression: Gene Rank *(APC)*, Order by: Overexpression: Gene Rank *(BRAF)*. A two-fold change, *P*-value corresponding to 1E^-4^ and top 10% gene rank were selected as threshold for this analysis. All statistical analyses containing mRNA reporters 215310_at (*APC*), 203525_s_at (*APC*) and 243829_at (*BRAF*) were directly exported from Oncomine.

### KMplot

(http://kmplot.com/analysis/index.php?p=service) is an online survival evaluation platform to enable the meta-analysis of a number of different datasets using gene expression data^34^. The correlation between overall survival and *APC/BRAF* mRNA expression can be assessed using mRNA – Start KM Plotter for pan-cancer, gene symbol-*APC/BRAF*, select all, select draw Kaplan-Meier plot.

### Statistical analysis

Comparison of *APC/BRAF* mRNA expression levels was directly obtained from Oncomine using a Student’s paired t-test. The prognostic data obtained from PrognoScan was selecting according to the calculated Cox *P*-values and corresponding Hazard ratios (95% confidence interval) for various endpoints (OS, DSS, DFS) that were visualized with a forest plot. For comparison of patient survival endpoints, patients were categorized according to survival and survival difference between high and low expression groups was determined by a log-rank test in PrognoScan. To analyse the prognostic value of *APC/BRAF* in the Kaplan-Meier plots, P-values from log-rank analyses were used to compare prognostic endpoints between patient cohorts using the KMplot webpage. False discovery rates below 5% and P < 0.05 were considered as statistically significant for all comparisons.

## Results

### Analysis of APC and BRAF gene mutations in CRC using CBioportal

We first examined the gene mutations and their frequency of occurrence as a function of colorectal cancer clinicopathologic parameters using seven clinical colorectal cancer studies (2,575 samples) in cBioportal^25–31^.

Across the seven studies surveyed in cBioportal, the percentage of samples in which somatic mutations in *APC* occurred was 69.6%, with a corresponding frequency of 13.7% for samples with *BRAF* mutations. In the case of *APC* mutations this corresponded to 2,452 driver mutations (2,483 of these were truncating mutations, 213 missense, 45 splice and 12 fusion) and for *BRAF*, 327 driver mutations (355 missense, 3 inframe, and 6 fusion). Most of the mutations located in the *APC* and *BRAF* protein sequences (**Figure 2A**) were annotated as oncogenic in cBioportal. The bar chart (**Figure 2B**) obtained from cBioPortal shows that the mutation frequency of *APC* is similar across the seven CRC studies selected, while in the case of *BRAF* a variation in frequency was observed across the studies examined. Significant variation was observed in the frequency (**Figure 2C**) and proportion of mutation types observed across the studies as a function of the tumour site (**Figure 2D**).

**Figure 1.**
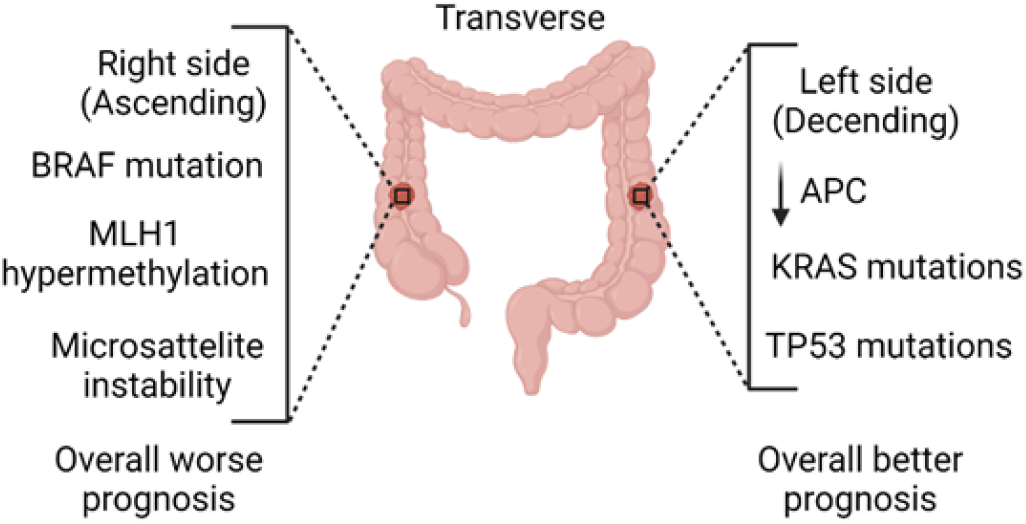
Genetic and biomolecular differences in colorectal cancer as a function of anatomical location. RCCs occur in the cecum, ascending colon and hepatic flexure. LCCs occur in the splenic flexure, descending, sigmoid and rectosigmoid colon.

**Figure 2.**
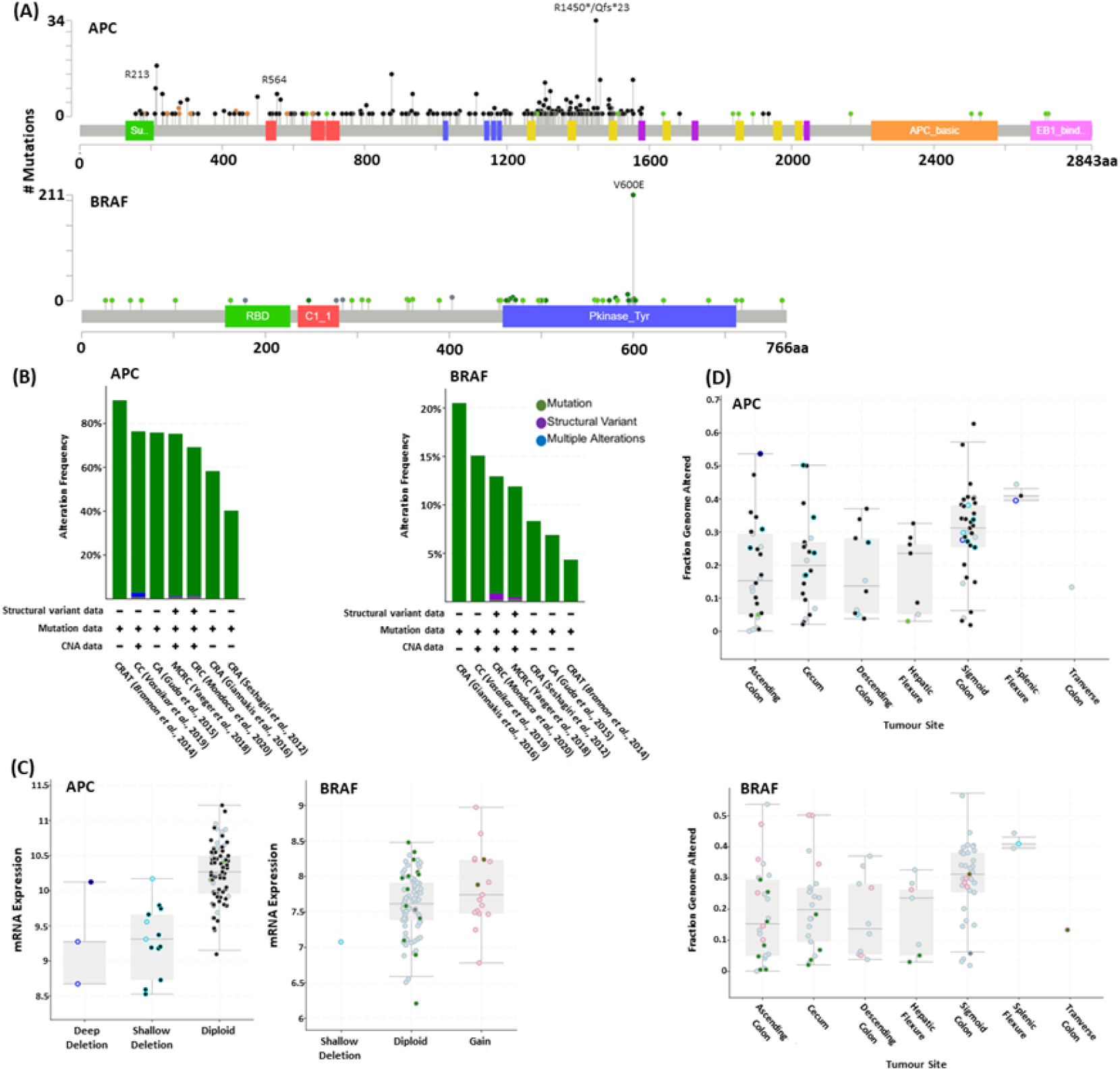
The distribution of APC and BRAF mutations occurring in their protein sequence across seven colorectal cancer studies. **(A)**, Lollipop diagram corresponding colorectal cancer studies selected in cBioportal and the frequency and types of changes occurring. Circles indicate sites in which the mutations occur, and the length of each lollipop represents the number of patients with the specific mutation **(B)**, mRNA expression levels and frequency of mutations **(C)** for APC and BRAF **(D)** and fraction of genome alterations as a function of anatomical site. *CRA, Colorectal Adenocarcinoma; CRC, Colorectal Cancer; MCRC, Metastatic Colorectal Cancer; CA, Colon Adenocarcinoma; CC, Colon Cancer; CRAT, Colorectal Adenocarcinoma Triplets.*

### *APC and BRAF* mRNA expression in healthy colon tissue and distinct types of CRC tissue representing three reporters (203525_s_at, 215310_at and 243829_at)

Using Oncomine, three studies (Skrzypczak et al., Kaiser et al. and Hong et al.) were analysed to compare the relative expression patterns of *APC* and *BRAF* mRNA expression levels in healthy colon and tumour tissue based on their anatomical location^35–37^.

Boxplots of *APC* (**Figure 3A-J**) mRNA expression levels obtained from Oncomine show a significant reduction of *APC* expression levels in distinct types of CRC tissue when compared to healthy colon tissue. Analyses of *APC* mRNA expression level patterns across three studies illustrates varied results when comparing different CRC tissue subtypes. In the case of *BRAF* (**Figure 3**K-L), mRNA expression levels are significantly elevated in CRC tissues when compared to healthy colon tissue. Corresponding fold-changes in expression levels and other study informationderived from Oncomine are summarized in Table 1.

**Table 1.** Combined APC and BRAF expression are frequently mutated in CRC upon comparison with healthy colon tissue. Fold-changes in APC and BRAF mRNA expression levels relative to matched healthy colon tissue as a function of CRC tumour subtypes as located in Oncomine.

| Gene | Colorectal Cancer Subtype | Reporter | Fold-change | P-value | N | Study |
| --- | --- | --- | --- | --- | --- | --- |
| APC | Colon Carcinoma | 203525_s_at | -6.093 | $1.13 \times 10^{-11}$ | 40 | <sup>35</sup> |
| APC | Colon Carcinoma | 215310_at | -1.230 | 0.036 | 40 | <sup>35</sup> |
| APC | Colon Carcinoma | 203525_s_at | -4.320 | $1.84 \times 10^{-10}$ | 40 | <sup>35</sup> |
| APC | Colon Carcinoma | 215310_at | -1.090 | 0.115 | 40 | <sup>35</sup> |
| APC | Colon Adenoma | 203525_s_at | -7.941 | $5.48 \times 10^{-7}$ | 40 | <sup>35</sup> |
| APC | Colon Adenoma | 215310_at | -1.418 | 0.003 | 40 | <sup>35</sup> |
| APC | Rectosigmoid Adenocarcinoma | 203525_s_at | -1.601 | $9.16 \times 10^{-5}$ | 105 | <sup>36</sup> |
| APC | Rectosigmoid Adenocarcinoma | 215310_at | -1.233 | 0.028 | 105 | <sup>36</sup> |
| APC | Colon Carcinoma | 203525_s_at | -2.166 | $1.14 \times 10^{-11}$ | 82 | <sup>37</sup> |
| APC | Colon Carcinoma | 215310_at | -1.211 | 0.228 | 82 | <sup>37</sup> |
| BRAF | Colon Carcinoma | 243829_at | 2.054 | $1.41 \times 10^{-7}$ | 40 | <sup>35</sup> |
| BRAF | Colon Carcinoma | 243829_at | 2.763 | $1.89 \times 10^{-6}$ | 40 | <sup>35</sup> |

| Gene | Colorectal Cancer Subtype | Reporter | Fold-change | P-value | N | Study |
| --- | --- | --- | --- | --- | --- | --- |
| APC | Colon Carcinoma | 203525_s_at | -6.093 | $1.13 \times 10^{-11}$ | 40 | 35 |
| APC | Colon Carcinoma | 215310_at | -1.230 | 0.036 | 40 | 35 |
| APC | Colon Carcinoma | 203525_s_at | -4.320 | $1.84 \times 10^{-10}$ | 40 | 35 |
| APC | Colon Carcinoma | 215310_at | -1.090 | 0.115 | 40 | 35 |
| APC | Colon Adenoma | 203525_s_at | -7.941 | $5.48 \times 10^{-7}$ | 40 | 35 |
| APC | Colon Adenoma | 215310_at | -1.418 | 0.003 | 40 | 35 |
| APC | Rectosigmoid Adenocarcinoma | 203525_s_at | -1.601 | $9.16 \times 10^{-5}$ | 105 | 36 |
| APC | Rectosigmoid Adenocarcinoma | 215310_at | -1.233 | 0.028 | 105 | 36 |
| APC | Colon Carcinoma | 203525_s_at | -2.166 | $1.14 \times 10^{-11}$ | 82 | 37 |
| APC | Colon Carcinoma | 215310_at | -1.211 | 0.228 | 82 | 37 |
| BRAF | Colon Carcinoma | 243829_at | 2.054 | $1.41 \times 10^{-7}$ | 40 | 35 |
| BRAF | Colon Carcinoma | 243829_at | 2.763 | $1.89 \times 10^{-6}$ | 40 | 35 |

**Figure 3.**
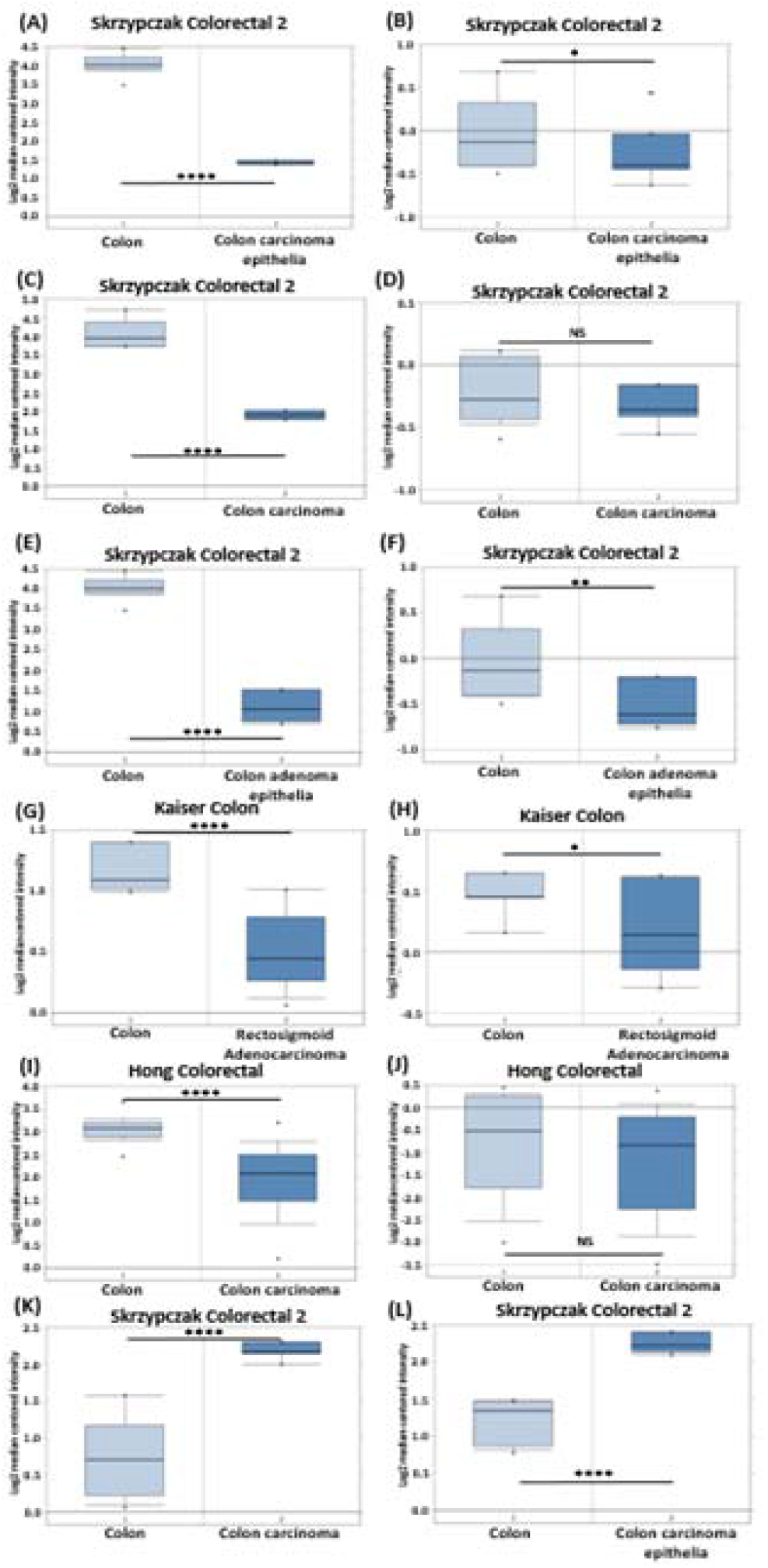
APC and BRAF mRNA expression levels in normal colon and colorectal cancer tissue^32^. Box plots (A, C, E, G and I) on the left side indicate 203525_s_at APC reporter. Box plots (B, D, F, H and J) on the right site represent 215310_at APC reporter. Box plots (K and L) down represent 243829_at BRAF reporter. All box plots are located from Oncomine searches. Corresponding datasets included: (A-F) Skrzypczak Colorectal 2 (n=40), (G and H) Kaiser Colon (n=105), (I and J) Hong Colorectal (n=82), (K and L) Skrzypczak Colorectal 2 (n=40). *P < 0.05, **P < 0.01, ***P < 0.001, ****P < 0.0001, NS > 0.05 as determined by a t-test..

The negative fold-change values under −2 together with *P* < 0.05 shows that *APC* gene expression is significantly reduced in CRC tumour tissue. In the case of colon carcinoma epithelia, colon carcinoma and colon adenoma epithelia a high negative fold-change is observed, confirming a reduction in *APC* mRNA expression levels in comparison with rectosigmoid adenocarcinoma. The P-value of reported fold-changes were found to be statistically significant (P<0.05), except for two cases (Skrzypczak and Hong colon carcinoma), which both applied to the 215310_at reporter. The positive fold-change (>2) in *BRAF* expression coupled with the significant *P*-values (P<0.05) confirms that the *BRAF* gene expression is significantly increased in colon carcinoma relative to healthy tissue.

### The prognostic value of *APC* and *BRAF* gene mutation

Pan-Cancer analysis of the prognostic value of *APC* and *BRAF* was performed in KMPlot (21 cancer studies) to explore the correlation between patient overall survival and the mRNA expression levels of these actionable genes.

Following analysis of pan-cancer patterns of mRNA expression for *APC* and *BRAF*, four datasets were analysed to explore their prognostic value in CRC. Meta-analysis of *APC* mRNA expression levels and their impact on specific endpoints (including disease-specific survival (DSS), disease-free survival (DFS) and overall survival (OS)) was performed and visualised using forest plots.

Pan-cancer analysis of the prognostic value of *APC* in KMplot, showed no direct association between *APC* levels and patient overall survival. In the case of breast (**Figure 4A**, HR 1.42), oesophageal (Figure 4B, HR 2.17), lung squamous cell carcinomas (**Figure 4E**, HR 1.5), and thyroid (**Figure 4G**, HR 2.91) higher *APC* levels were associated with poor overall survival. However, for head and neck squamous cell carcinoma (**Figure 4C**, HR 0), renal cell carcinoma (**Figure 4D**, HR 0.72), and ovarian (**Figure 4F**, HR 0.43) cancer higher *APC* expression levels were associated with more favourable patient overall survival outcomes.

**Figure 4.**
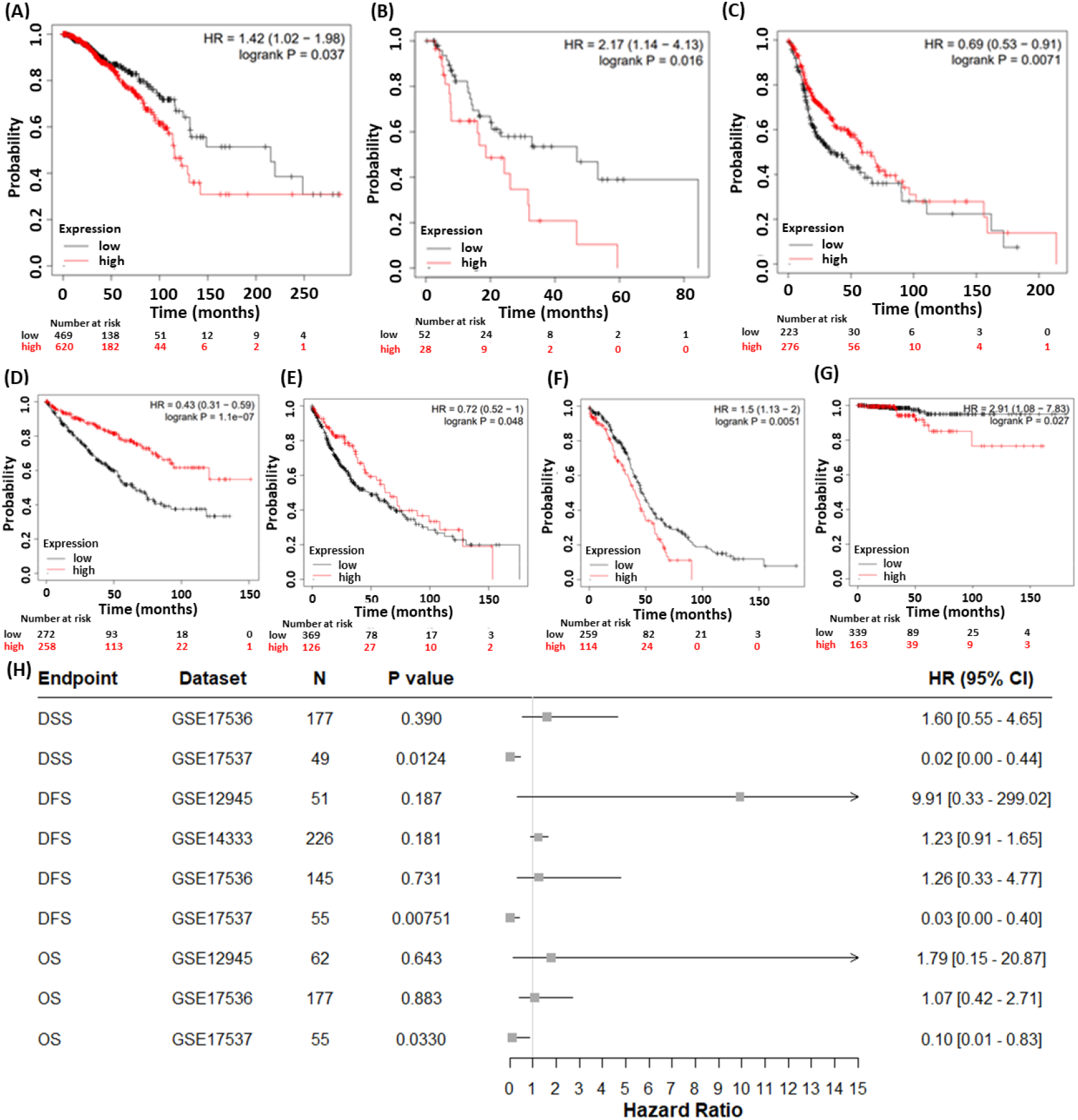
Pan-cancer Kaplan-Meier plots of overall survival correlated with APC mRNA expression levels. Breast cancer (n=1090, FDR >50) ***(A**), esophageal adenocarcinoma (n=80, FDR=50) **(B**)*, head-neck squamous cell carcinoma (n=500, FDR >50) ***(C)***, renal cell carcinoma (n=530, FDR=1) ***(D)***, lung squamous cell carcinoma (n=501, FDR >50) ***(E)***, and ovarian cancer (n=374, FDR >50) **(F)**, and thyroid cancer (n= 502, FDR >50%) **(G)**. Forest plot showing the prognostic value of APC across multiple datasets in PrognoScan. Prognostic endpoints investigated are classified as disease-specific survival (DSS), disease-free survival (DFS) and overall survival (OS) for APC 215310_at reporter ***(H)***. Grey squares represent the point estimate of the hazard ratio (HR). FDR: False Discovery Rate and HR: Hazard Ratio.

Next, we examined the prognostic value of *APC* (*APC* 215310_at and 203525_s_at reporters) in CRC using PrognoScan. Analysis of PrognoScan yielded four datasets that were subsequently applied to determining the prognostic value of *APC* and *BRAF* in CRC across three survival endpoints (DSS, DFS and OS). Except for the GSE17537 dataset (all endpoints), no statistically significant associations were observed between *APC* expression and patient survival endpoints.

Pan-cancer analysis of *BRAF* prognostic value in KMplot, showed no consistent association between *BRAF* mRNA expression levels and patient overall survival across all cancer types studied. In the case of breast (**Figure 5A**, HR 0.67), head and neck squamous cell carcinoma (**Figure 5B**, HR 0.72), renal cell carcinomas (**Figure 5C**, HR 0.56) high *BRAF* expression levels were associated with better patient overall survival. Conversely, for hepatocellular carcinoma (**Figure 5D**, HR 1.5), ovarian cancer (**Figure 5E**, HR 1.33), and endometrial carcinoma (**Figure 5F**, HR 2.14) higher *BRAF* expression levels were associated with poor patient overall survival.

**Figure 5.**
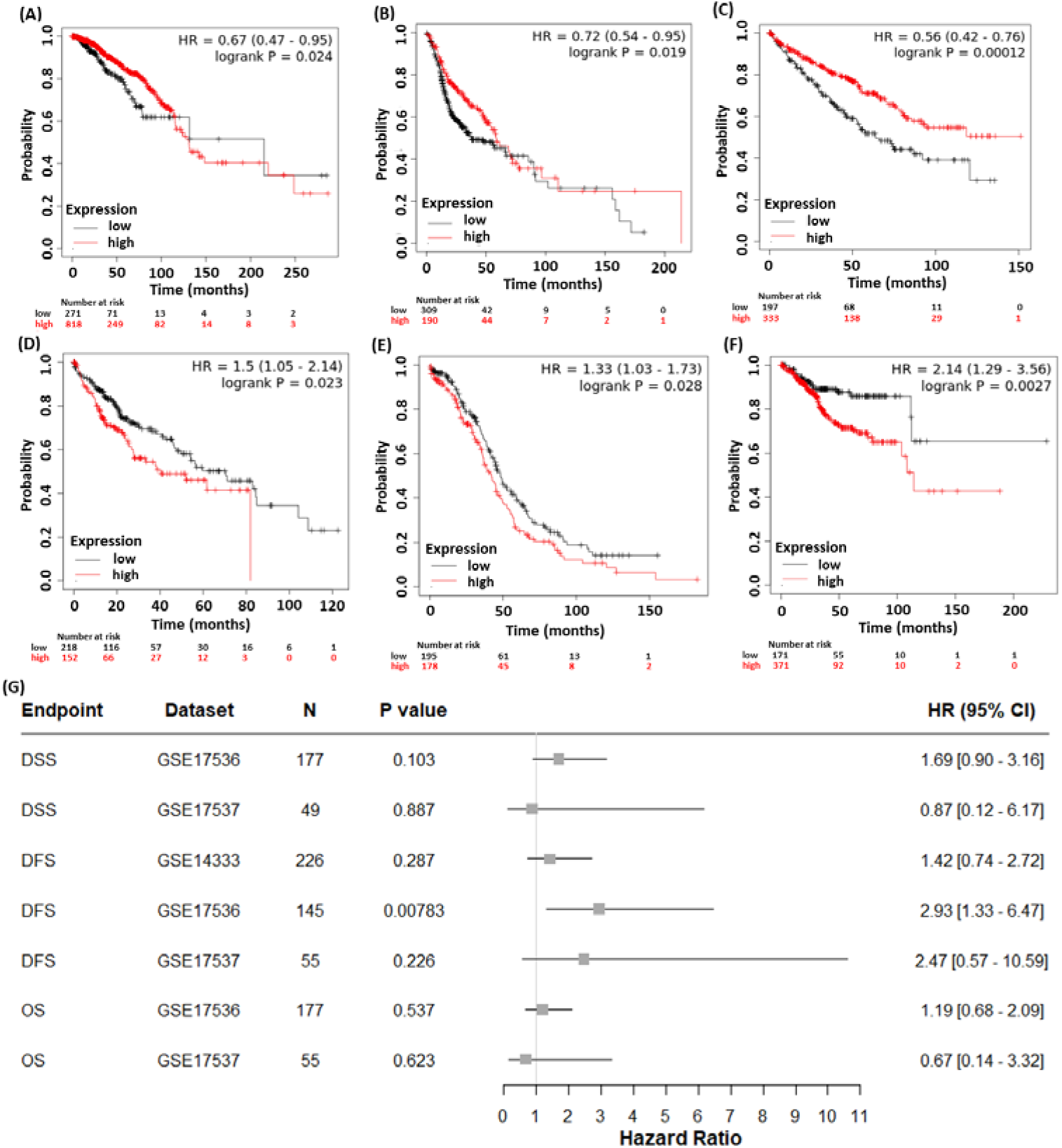
Pan-cancer Kaplan-Meier plots for BRAF mRNA expression level impact on patient survival endpoints. Breast cancer (n=1,090, FDR >50%) **(A)**, head-neck squamous cell carcinoma (n=500, FDR >50%) **(B)**, kidney renal cell carcinoma (n-530, FDR=3%) **(C)**, hepatocellular carcinoma (n=371, FDR >50%) **(D)**, ovarian cancer (n=374, FDR >50%) **(E)**, and endometrial carcinoma (n=543, FDR >50%) **(F)**. Forest plot showing the prognostic value of BRAF across different datasets. Prognostic value classified as disease-specific survival (DSS), disease-free survival (DFS) and overall survival (OS). Grey squares represent the point estimate of the hazard ratio (HR). Solid lines represent 95% confidence intervals (CI). The grey line oriented at 1 represents null or no difference. Data represents the 243829_at reporter **(G)**. FDR: False discovery rate

Using Prognoscan we identified four CRC datasets that were subsequently applied to determining the prognostic value of *BRAF* in CRC across three survival endpoints (disease-specific survival-DSS, disease free survival-DFS and overall survival-OS). Except for GSE17536 for the disease-free survival endpoint, no statistically significant associations were observed between *BRAF* expression and survival endpoints.

## Discussion

In this study we investigated the evidence base for *APC* and *BRAF* expression patterns, frequency of mutations and prognostic role using a combination of open-source bioinformatics tools and clinical datasets. Our rationale for selecting these genes for the present study was informed by the established link between their mutation and CRC tumour aggressiveness^38–41^.

Here, we report an accessible bioinformatics pipeline for performing a meta-analysis using freely available online bioinformatics resources (cBioPortal, Prognoscan, Oncomine, KMPlot) to interrogate existing clinical data on *APC* and *BRAF* as actionable drivers of cancer aggressiveness in patients with CRC. The implementation of this approach has several advantages, such as refinement and replacement of pre-clinical models for studying the role and significance of these mutations in cancer progression. Developing a deeper understanding of the frequency of mutations, their clinical significance and impact on patient prognosis provides a strong foundation for the careful design of pre-clinical studies examining the role of these mutations in response to novel experimental therapeutics. Moreover, analysis of large patient clinical datasets increases the translational relevance of findings from these studies to humans through analysing pre-existing clinical data. To our knowledge this study is the first report of the application of bioinformatics approaches to the evaluation of the expression and prognostic value of *APC* and *BRAF* in CRC.

The model genes *APC* and *BRAF* were selected to demonstrate the utility of this presented pipeline. *APC* and *BRAF* mutation assessment have been useful and expedient in prognosis of CRC and their relative expression levels and mutational status have been implicated in treatment response^7,9^. In the present study our analysis of existing clinical datasets shows a significant reduction in *APC* mRNA expression levels that occurs in distinct anatomical regions of colon tumour tissue when compared to healthy colon tissue (Figure 3). *APC* is a well-recognized tumour suppressor gene which is highly mutated in CRC. Our findings of frequent mutations and downregulation of *APC* expression levels are consistent with previous reports that show the role of *APC* loss or gain of function in CRC tumorigenesis^38^. The loss of *APC* tumour suppressive functions and gain of function from truncated *APC* are widely recognized to contribute to the initiation, progression, and maintenance of CRC^7,9,28^. Our analysis of the expression of *BRAF*, demonstrated a significant increase in *BRAF* expression in CRC tissues relative to healthy tissue. Also, *BRAF* mutations function as strong negative prognostic markers, relating to the V600E (valine to glutamate at codon 600) mutation that leads to gain of function. These mutations have previously been observed in mismatch repair deficient tumours and right sided colon tumours in older women ^42–44^. Co-occurrence of *BRAF* and *KRAS* mutations has frequently been implicated in poor prognosis for CRC patients and a marker of resistance to chemotherapy^45^. Our results indicate that the prognostic value of *BRAF* mutations is not unique to CRC but has also been observed in kidney renal cell carcinoma. However, previous research implies that patients with renal cell carcinoma lack the presence of *BRAF* mutations, as opposed to patients with CRC^46,47^. According to the literature *BRAF* mutations occur frequently in melanoma, ovarian and thyroid cancer^48^.

Following our analyses of gene mutation signatures and expression levels for *APC* and *BRAF*, we examined the prognostic value of these genes (Figure 4-5). Our findings show that loss of *APC* and gain of *BRAF* both contribute to poorer survival endpoints in patients with CRC. These findings are consistent with previous reports.

A limitation of the present study is a lack of stratification of datasets according to patient metadata, clinicopathologic parameters and anatomical location of CRC tumours. The biology underlying left and right sided CRC has been associated with differences in tumour pathology which vary according to patient parameters (e.g. age, sex, lifestyle factors)^49^. The prognostic value of *APC* and *BRAF* expression and their role in CRC tumorigenesis has previously been associated with patient sex and location of the CRC tumour, which has implications for establishing the clinical significance of these actionable mutations according to tumour location. For example, BRAF mutations are typically associated with right-sided colon cancer.

Further investigation of the genetic and phenotypic role of *APC* and *BRAF* genes can contribute to the identification of new biomarkers or the development of novel potential drug targets or combinations strategies for overcoming chemoresistance, with the lack of requirement for the use of animal models.

The rapid rise in biobanks and online bioinformatic tools, coupled with the increasing the number of parameters available in these biobanks and software can improve future functional analysis and reduce the need for animal models. The pipeline developed in this study can be directly applied to the evaluation of the expression and prognostic value of other actionable genes in cancer.

## Conclusion

In this study we reported analysis of both APC and BRAF mutations, expression profiles and prognostic roles as actionable genes in CRC. Detailed analysis of genomic alterations in CRC can provide novel insights into disease progression, supporting the development of novel drug treatment combinations that will ultimately improve patient outcomes. Findings from this study demonstrate the importance of accessibility to clinical datasets and a pipeline for investigating the role of genes in the diagnosis and therapy of CRC.

## Data Availability

All data produced in the present study are available upon reasonable request to the authors.

