## Supplemental for "Development of an accessible gene expression bioinformatics pipeline to study driver mutations of colorectal cancer"

**Supplemental Material**

Web links to used software:

**Table S1-** Software used in the present study and associated web links

| Software | Hyperlinks | Used for: |
| --- | --- | --- |
| RStudio (version 1.4.1106) | https://www.rstudio.com/ | R environment |
| R (version 4.1.1), package: forestplot | <https://www.r-project.org/>, https://cran.r-project.org/web/packages/forestplot/ | Forest plots |
| cBioPortal | <https://www.cbioportal.org/> | Common gene mutations analysis in CRC |
| Oncomine | <https://www.oncomine.org/> | Gene expression pattern analysis |
| Prognoscan | <http://dna00.bio.kyutech.ac.jp/PrognoScan/> | Prognostic analysis |
| KmPlot | <https://kmplot.com/analysis/> | Pan-cancer analysis |

R code used to compute forestplot of Figure X. The same code with appropriate values were used for Figure 1B and Figure 3.

#Install package 'forestplot' if needed.

install.packages**(**'foresplot'**)**

library**(**forestplot**)**

\# Cochrane data from the 'rmeta'-package

cochrane_from_rmeta **<-**

structure**(**list**(**

mean **=** c**(NA**, **NA**, 1.60, 0.02, 9.91, 1.23, 1.26, 0.03, 1.79, 1.07, 0.10**)**,

lower **=** c**(NA**, **NA**, 0.55, 0.00, 0.33, 0.91, 0.33, 0.00, 0.15, 0.42, 0.01**)**,

upper **=** c**(NA**, **NA**, 4.65, 0.44, 299.02, 1.65, 4.77, 0.40, 20.87, 2.71, 0.83**))**,

.Names **=** c**(**"mean", "lower", "upper"**)**,

row.names **=** c**(NA**, **-**11L**)**,

class **=** "data.frame"**)**

texttable **<-** cbind**(**

c**(**"", "Endpoint", "DSS", "DSS", "DFS", "DFS", "DFS", "DFS", "OS", "OS", "OS"**)**,

c**(**"", "Dataset", "GSE17536", "GSE17537", "GSE12945", "GSE14333", "GSE17536", "GSE17537", "GSE12945", "GSE17536", "GSE17537"**)**,

c**(**"", "N", "177", "49", "51", "226", "145", "55", "62", "177", "55"**)**,

c**(**"", "P value", "0.390", "0.0124", "0.187", "0.181", "0.731", "0.00751", "0.643", "0.883", "0.0330"**)**,

c**(**"", "HR (95% CI)", "1.60 [0.55 - 4.65]", "0.02 [0.00 - 0.44]", "9.91 [0.33 - 299.02]", "1.23 [0.91 - 1.65]", "1.26 [0.33 - 4.77]", "0.03 [0.00 - 0.40]", "1.79 [0.15 - 20.87]", "1.07 [0.42 - 2.71]", "0.10 [0.01 - 0.83]"**))**

forestplot**(**texttable,

graph.pos **=** 5,

zero **=** 1,

boxsize **=** 0.2,

graphwidth **=** unit**(**8,"cm"**)**,

txt_gp **=** fpTxtGp**(**ticks**=**gpar**(**cex**=**1**)**, xlab**=**gpar**(**cex**=**1, fontface**=**"bold"**))**,

xlab **=** "Hazard Ratio",

hrzl_lines **=** list**(**"3" **=** gpar**(**lty **=** 1**))**,

cochrane_from_rmeta,new_page **=** **TRUE**,

is.summary **=** c**(TRUE**,**TRUE**,rep**(FALSE**,9**)**,**TRUE)**,

clip**=** c**(-**5.0,15.0**)**,

col **=** fpColors**(**box **=** "darkgrey",

line **=** "black"**))**

**Table S2: Supplementary information corresponding to Figure 3**

| Cancer type | N | HR (95% CI) | FDR (%) |
| --- | --- | --- | --- |
| Breast cancer | 1090 | 1.42 (1.02 – 1.98) | Over 50 |
| Esophageal adenocarcinoma | 80 | 2.17 (1.14 – 4.13) | 50 |
| Head-neck squamous cell carcinoma | 500 | 0.69 (0.53 – 0.91) | Over 50 |
| Kidney renal cell carcinoma | 530 | 0.43 (0.31 – 0.59) | 1 |
| Lung squamous cell carcinoma | 501 | 0.72 (0.52 – 1) | Over 50 |
| Ovarian cancer | 374 | 1.5 (1.13 – 2) | Over 50 |
| Thyroid carcinoma | 502 | 2.91 (1.08 – 7.83) | Over 50 |

*Adjust table style

**Table S3: Supplementary information corresponding to Figure 6**

| Cancer type | N | HR (95% CI) | FDR (%) |
| --- | --- | --- | --- |
| Breast cancer | 1090 | 0.67 (0.47 – 0.95) | Over 50 |
| Head-neck squamous cell carcinoma | 500 | 0.72 (0.54 – 0.95) | Over 50 |
| Kidney renal cell carcinoma | 530 | 0.54 (0.42 – 0.76) | 3 |
| Liver hepatocellular carcinoma | 371 | 1.5 (1.05 – 2.14) | Over 50 |
| Ovarian cancer | 374 | 1.33 (1.03 – 1.73) | Over 50 |
| Uterine corpus endometrial carcinoma | 543 | 2.14 (1.29 – 3.56) | 50 |

*Adjust table style
